# COVID-19 Vaccination Priority Evaluation

**DOI:** 10.1101/2021.02.27.21252569

**Authors:** Jozo Dujmović, Daniel Tomasevich

## Abstract

Computing the COVID-19 vaccination priority is an urgent and ubiquitous decision problem. In this paper we propose a solution of this problem using the LSP evaluation method. Our goal is to develop a justifiable and explainable quantitative criterion for computing a vaccination priority degree for each individual in a population. Performing vaccination in the order of the decreasing vaccination priority produces maximum positive medical, social, and ethical effects for the whole population. The presented method can be expanded and refined using additional medical and social conditions. In addition, the same methodology is suitable for solving other similar medical priority decision problems, such as priorities for organ transplants.

## 1. Introduction

The concept of vaccination priority is essential for successful efforts to fight contagious diseases. It is a simple “first things first” concept exemplified by conditions such as “old people should be vaccinated before young people,” “residents of long-term care facilities should be vaccinated first,” “medical personnel should be protected before all other professions,” etc. A comprehensive analysis of vaccine allocation for COVID-19, performed by the CDC COVID-19 Response Team and the Advisory Committee on Immunization Practices (ACIP) presented in [1-4] and summarized in [5] is based on four criteria: (1) risk of acquiring infection, (2) risk of severe morbidity and mortality, (3) risk of negative societal impact, and (4) risk of transmitting infection to others. Each population group is classified as low/medium/high risk level in each of the four criteria, and without quantification distributed in four vaccine allocation phases according to decreasing estimated total risk. The resulting phased approach identifies four phases of allocation [5]. In the Phase 1a vaccination is given to high risk health workers and first responders, as well as residents of long-term care facilities. The subsequent Phase 1b includes persons aged above 75 years, and frontline non-health care essential workers, and Phase 1c includes persons aged 16-64 years with medical conditions that put them at high risk, persons aged 65-74 years and remaining essential workers not recommended in Phase 1b. Phase 2 includes K-12 teachers, older adults not included in Phase 1, homeless people, people with disabilities, and critical workers in high risk settings. Phase 3 includes children and young adults and workers with risk of exposure lower than previous Phases 1 and 2. Finally, Phase 4 includes everybody else not included in previous Phases 1, 2, and 3. As anticipated in [5], this phased method can be subsequently updated, and that is done at the state level [6-8] and then implemented by health maintenance organizations (e.g. [9]). This approach is consistent with the recommendations resulting from mathematical models of the dynamics of pandemic expansion [10-12].

The presented phased method is logical, justifiable, and acceptable in the form of qualitative recommendations. In addition, imprecise qualitative phased allocation criteria offer a political benefit that it is easy to defend deviations and exceptions in vaccination activities caused by organizational or financial problems. On the other hand, it is easy to see that the phased vaccine allocation model has the following provable drawbacks: (1) a coarse discretization of phases (2) the absence of priority differentiation inside individual phases, (3) the ambiguity of phase belonging (at all borders between phases), (4) undefined duration of individual phases and (5) incompleteness (absence of non-medical criteria). These drawbacks are discussed below. Our goal is to show that the drawbacks of the phased model can be eliminated using quantitative decision methods.

Each discretization is caused by the absence of a continuous model. In practice, continuous models are equivalent to discrete models with very fine granularity. However, having only 2-3 discrete phases or tiers is too coarse. With coarse granularity, too many people belong in the same group and it is immediately evident that they should be differentiated. The need for precision higher than the precision of the basic phased model is visible in [8] where the California Department of Public Health provided seven rather detailed recommendations for subprioritization during the Phase 1a. Such recommendations are the necessary refinements of the basic phased model, developed with intention to increase precision and bring it at the level that is necessary for practical allocation of scarce vaccines.

Phased criteria include oversimplifications that people cannot accept without objecting. All individuals that are highest ranked in a given phase face two problems: (1) they have the same priority as people that are the lowest ranked in their phase, what is obviously unfair, and (2) they are indiscernible with people at the end of the previous phase, but they still must wait for the complete termination of the previous phase before they become eligible for vaccination. For example, the usual threshold criterion of priority vaccination of people over 75 years of age is an unnecessary discontinuity, that cannot explain to those who are 74.5 years old why they must wait much longer for their chance to be vaccinated regardless of practically indistinguishable difference in age between 74.5 and 75. Furthermore, it seems to be no proof that the threshold of 75 years is better than the threshold of 74 or 76 years.

The phased vaccination model does not offer strict guidelines about the duration of each phase, or what are the conditions for declaring the end of a specific phase and the beginning of the next phase, with or without overlap. It is unavoidable that such decisions can be predominantly administrative, territorially inconsistent, insufficiently justified, and criticized to be too early, or too late. In addition, the phased model does not cover the problems of micromanagement and scheduling of the vaccination process.

The problem of incompleteness is visible in the absence of criteria that explicitly take into account the role and responsibility of an individual. For example, according to the phased model, a perfectly healthy member of the U.S. Congress, 29 years old, would be vaccinated in the very last phase. The same holds for all other young people in leading positions in industry or government. If it is publicly visible that political and other leaders have justifiable priority in COVID-19 vaccination, then such a priority should be explicitly included in the vaccine allocation criteria that are developed for the whole population. Currently, that is not the case.

All the above problems can be eliminated using quantitative decision models. Three obvious questions related to the incompleteness and imprecision of the phased model are the following:

1. Is it possible to make precise and complete quantitative decision model for vaccine allocation, reflecting the same medical guidelines used in the phased model?
2. Can the quantification and precision improvement be achieved with a modest effort?
3. Is the quantification yielding more benefits than the phased method?

The goal of this paper is to provide affirmative answers to these three questions. We propose a quantitative model for computing the vaccination priority degree for each individual, based on the LSP decision method [13]. The presented model is consistent with concepts and methodology developed by CDC and related governmental and medical organizations.

The rest of this paper is organized as follows. In Section 2 we introduce the vaccination priority degree and its use in organizing the vaccination process. Section 3 presents a set of 21 attributes that are used for computing the vaccination priority degree. Attribute criteria are developed in Section 4 and their aggregation structure in Section 5. Examples of application of the proposed method are presented in Section 6, and Section 7 contains conclusions.

## 2. The vaccination priority degree and its use

Let *P* denote a vaccination priority degree as a numeric indicator in the range [0,1] (for convenience, *P* can be multiplied by 100 and expressed in the range [0, 100%]). Suppose that a population of *n* individuals has sorted vaccination priority degrees. 1 ≥ *P*_1_ > *P*_2_ > ⋯ > *P*_*n*_ > 0. The priority degree *P*_*i*_ denotes the expected benefit for the whole population obtained by vaccinating the *i*^*th*^ individual.

If the number of available vaccines *k* is less than *n* then the maximum overall cumulative benefit is obtained by vaccinating *k* individuals according to the decreasing priority degree, starting from the highest priority. If the organizer of vaccination (a government and/or a medical organization) knows the sequence of priority degrees *P*_1_ > *P*_2_ > ⋯ > *P*_*n*_, and these values are known also to each individual (or “patient”), then it is easy to solve several important practical problems: (1) the dynamics of supply of vaccines, (2) the number of needed medical personnel who give vaccines, (3) the size and the working schedule of facilities that provide vaccination, (4) the total time to protect the whole population, (5) the cost of vaccination, and (6) the schedule of vaccination.

The optimum schedule of vaccination can be based on the announcement of the “daily priority threshold (DPT),” e.g., “on 4/19/2021 we will vaccinate patients who have DPT=81% or more.” Obviously, if we can vaccinate *k* people per day, and we already vaccinated *m* people, then from the sequence *P*_*m*+1_ > *P*_*m*+2_ > ⋯ > *P*_*m*+*k*_ we can use *DPT* = *P*_*m*+*k*_, and eliminate long waiting lines, the patient anxiety, and the waste of time. All precise planning activities can be completed any time before the beginning of the actual vaccination process.

The success of the vaccination process depends on the availability of precisely computed sequence of priority degrees. In this paper, we compute the priority degrees using the LSP method [13]. This method consists of three major steps. The first step is the definition of the vaccination priority attribute tree, i.e. the identification of all medical, social, ethical, and political factors that affect the vaccination priority degree. In the second step we define attribute criteria, i.e. functions that compute individual attribute priority degree as a value in the selected range [0, 100%]. In the third step we create a logic aggregation structure that performs fusion of all attribute priority degrees, and computes the overall vaccination priority degree *P* for each specific patient. In subsequent sections we present these three steps, followed by examples, discussion, and conclusions.

## 3. The vaccination priority attribute tree

The identification of vaccination priority attributes is a process based on systematic decomposition of groups of attributes, as shown in Fig. 1. The basic goal of the LSP method is to include all relevant attributes, i.e. all attributes that affect the vaccination priority of each person. The attributes should be nonredundant and complete. If the attributes are redundant then the same property can be expressed using several scattered attributes and in such a case it is difficult to correctly estimate the total impact of the property and select appropriate weights of redundant components. The need for completeness is obvious: the priority degree cannot be reliable if it is only based on a fraction of attributes that affect the priority.

**Fig. 1.**
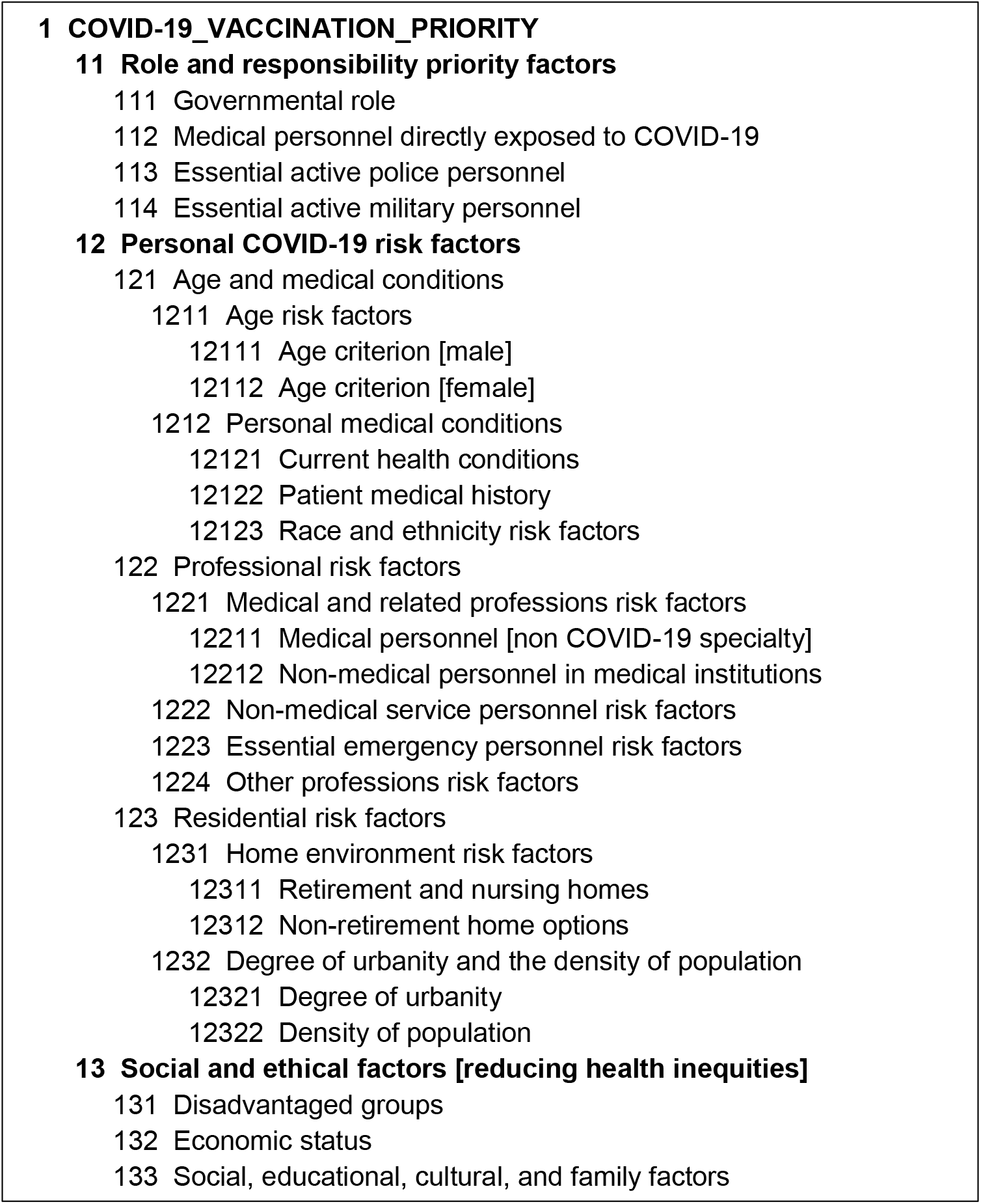
COVID-19 vaccination priority attribute tree

The root of the tree is the node #1. At the initial root level we identified three groups of attributes, denoted #11, #12, and #13. The first group of attributes (#11) includes priority factors based on the role and responsibility of each individual person. For example, if the analyzed person has high responsibility for critical decision making or critical activities in government, police or military, such a person should have high vaccination priority because s/he affects the activities and lives of many other people. Similarly, all medical personnel in permanent direct contact with COVID-19 patients deserve the same highest vaccination priority. The priority factors based on the role and responsibility do not depend on the age, gender, race, or medical conditions of the specific individual.

The second group of attributes (#12) includes general risk factors identified by CDC and other governmental organizations, primarily the factors based on the age and personal medical conditions. We also consider various risks based on professional work. Such professions are those that cannot be reduced to online work from home. Included are all personnel in medical institutions who work with non-COVID-19 patients, as well as personnel in service industries, such as restaurants, hotels, food stores, face-to-face teachers, and various workers in emergency services. This group also includes the subgroup of home environment risk factors, such as life in retirement home, large families and locations with high urbanity and density of population.

The third group of attributes (#13) includes social and ethical priority attributes. It includes attributes related to disadvantaged groups, family status, minorities that need protection, and groups with the low economic and/or educational status.

In this way we created the list of 21 input attributes shown in Fig. 2, and each person can be characterized using 21 values of these attributes. The next step is to develop the attribute criteria, which are functions used to compute the individual attribute priority degrees. Then, we can aggregate the individual priority degrees, and compute the overall vaccination priority degree.

**Fig. 2.**
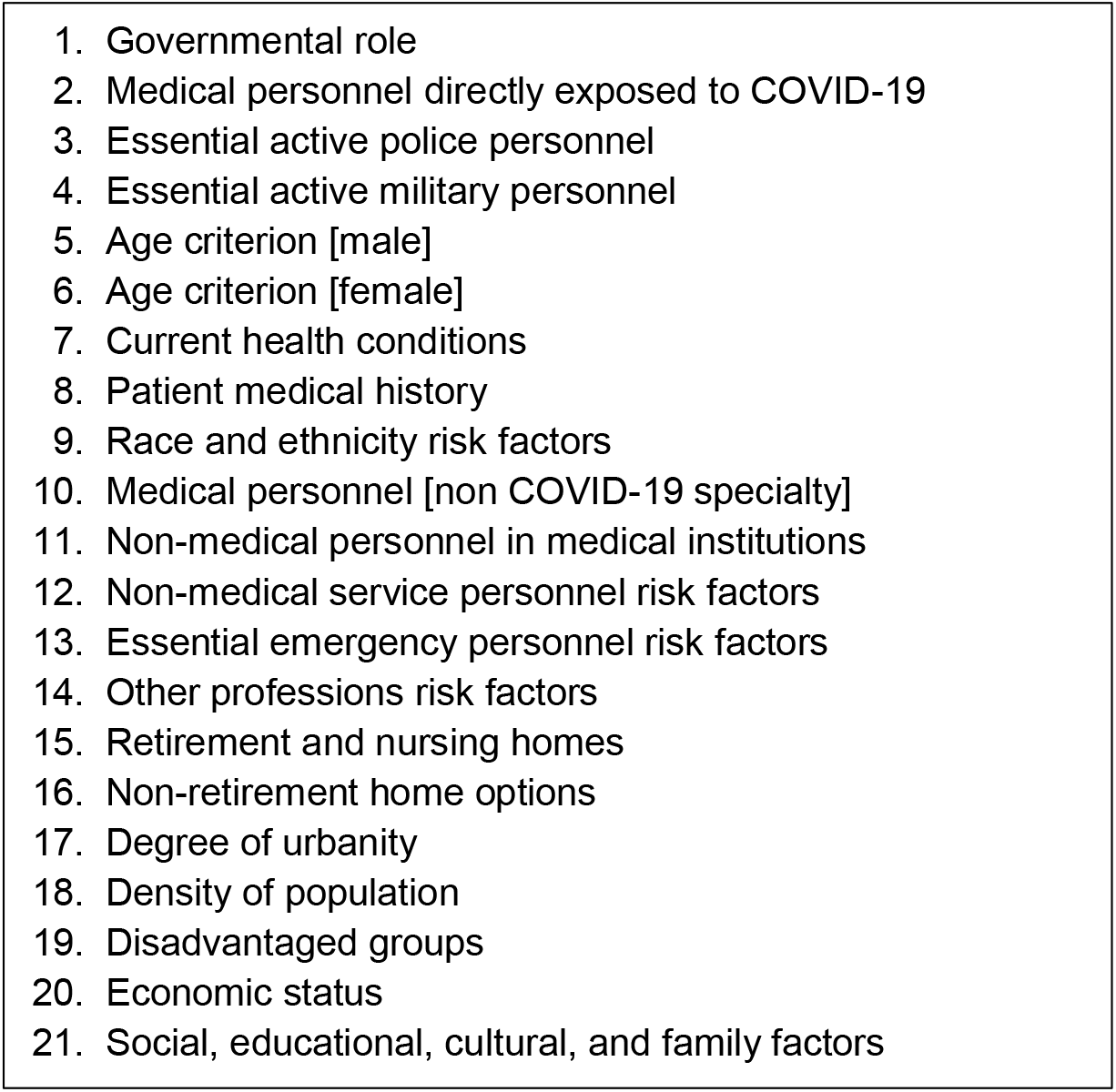
List of input attributes that affect the COVID-19 vaccination priority

## 4. Vaccination priority attribute criteria

Let *a*_1_, ⋯, *a*_*m*_ be a set of *m* priority attributes (in our case, *m* = 21). For each attribute we must create an attribute criterion for computing the attribute priority contributions *p*_*i*_ = *g*_*i*_(*a*_*i*_), 0 ≤ *p*_*i*_ ≤ 1, *i* = 1, ⋯, *m*. The range of contribution is from no contribution *p*_*i*_ = 0 to the highest possible contribution *p*_*i*_ = 1 (or 100%). Then, the overall vaccination priority degree of a person is computed as a graded logic function of the attribute priority degrees: *P* = *L*(*p*_1_, ⋯, *p*_*m*_).

The attribute criteria *g*_*i*_: *ℝ* → [0,1] can take many different forms [13] and in the simplest case they are approximated using piecewise linear approximations, as exemplified by the criterion for age priority shown in Fig. 3. A compact vertex notation of a piecewise linear criterion is based on a set of breakpoints: *Crit*(*a*[*years*]) = {(0,0), (25,10), (75,100)}.

**Fig. 3.**
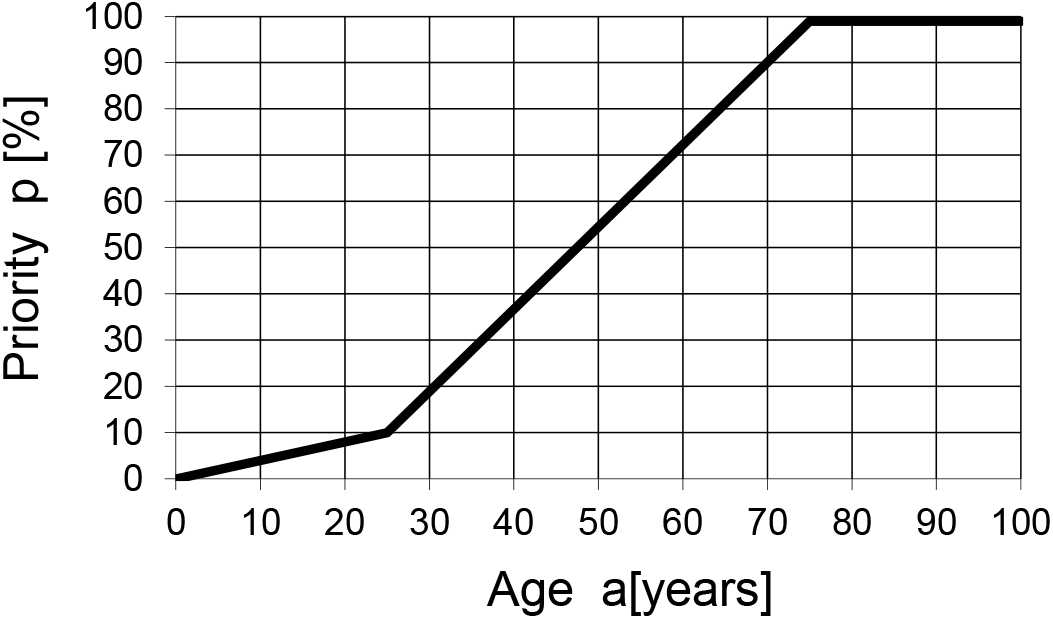
Sample age priority criterion

In a general case of *k* breakpoints *Crit*(*a*) = {(*a*_1_, *p*_1_), (*a*_2_, *p*_2_), ⋯, (*a*_*k*_, *p*_*k*_)} we use the following assumptions: *a*_1_ < *a*_2_ < ⋯ < *a*_*k*_, ∀*a* < *a*_1_ ⇒ *p* = *p*_1_, and ∀*a* > *a*_*k*_ ⇒ *p* = *p*_*k*_. Between adjacent breakpoints (*a*_*i*_, *p*_*i*_), (*a*_*i*+1_, *p*_*i*+1_) we use linear interpolation; for example, according to Fig. 3, if *a* = 50, then *p* = 55%. The set of 21 COVID-19 attribute criteria is presented in Table 1. For each attribute criterion we define a table of breakpoints with increasing values of arguments, and a short description that explains the concept of attribute evaluation.

**Table 1.** COVID-19 attribute criteria

| **111** |  | **Governmental role** |
| --- | --- | --- |
| Value | % | This criterion includes people of any age who have roles that are indispensable for functioning of society and directly or indirectly affect the lives of the majority of people. Evaluated using decreasing 5% significance rank steps:<br>1 = The top rank (100%) [score = $100-5*(rank-1)$]<br>2 = Next to the top rank (score = 95%), etc.<br>21 = Not in this group |
| 1 | 100 |  |
| 21 | 0 |  |
| **112** |  | **Medical personnel directly exposed to COVID-19** |
| Value | % | This criterion includes medical personnel (doctors, nurses, and supporting health workers) who are in the permanent contact with covid-19 patients in ICU and elsewhere. Evaluated using decreasing 5% risk rank steps:<br>1 = The highest risk (100%) [score = $100-5(rank-1)$]<br>2 = Next to the highest risk (score = 95%), etc.<br>21 = Not in this group |
| 1 | 100 |  |
| 21 | 0 |  |
| **113** |  | **Essential active police personnel** |
| Value | % | This group includes police officers who are in frequent close contact with large random groups of people, as well as police officers in high command roles. Evaluated using decreasing 5% responsibility/replaceability rank steps:<br>1 = The highest responsibility (100%) [score = $100-5(rank-1)$]<br>2 = Next to the highest responsibility (score = 95%), etc.<br>21 = Not in this group |
| 1 | 100 |  |
| 21 | 0 |  |
| **114** |  | **Essential active military personnel** |
| Value | % | This group includes military command officers high responsibility roles, as well as special units with highly trained individuals in situations where they cannot be replaced. Evaluated using decreasing 5% responsibility/replaceability rank steps:<br>1 = The highest responsibility (100%) [score = $100-5(rank-1)$]<br>2 = Next to the highest responsibility (score = 95%), etc.<br>21 = Not in this group |
| 1 | 100 |  |
| 21 | 0 |  |
| **12111** |  | **Age [male]** |
| Value | % | Age risk is evaluated using the following criterion<br>$Crit(age) = \{(0,0), (A, 10), (B, 100)\}$. A and B are adjustable parameters.<br>Sample breakpoints are:<br>A = 25<br>B = 75 |
| 0 | 0 |  |
| 25 | 10 |  |
| 75 | 100 |  |
| **12112** |  | **Age [female]** |
| Value | % | Age risk is evaluated using the following criterion<br>$Crit(age) = \{(0,0), (A, 10), (B, 100)\}$. A and B are adjustable parameters.<br>Sample breakpoints are:<br>A = 20<br>B = 70 |
| 0 | 0 |  |
| 20 | 10 |  |
| 70 | 100 |  |
| **12121** |  | **Current health conditions** |
| Value | % | Health conditions are evaluated using the following rating scale:<br>0 = excellent, 1 = good, 2 = average, 3 = poor, 4 = very poor |
| 0 | 0 |  |
| 4 | 100 |  |
| **12122** |  | **Patient medical history** |
| Value | % | Patient medical history (diseases, surgeries, children, etc.) is evaluated using the following concern rating scale:<br>0 = no concern, 1 = low concern, 2 = medium concern, 3 = high concern, 4 = very high concern |
| 0 | 0 |  |
| 4 | 100 |  |
| **12123** |  | **Race and ethnicity risk factors** |
| Value | % | Evaluated using the following risk rating scale:<br>0 = no risk; 1 = low; 2 = medium; 3 = high; 4 = very high<br>(Indigenous people, Latinos and African Americans are disproportionately represented among patients who contract COVID-19 and die from the disease). |
| 0 | 0 |  |
| 4 | 100 |  |
| **12211** |  | **Medical personnel [non COVID-19 specialty]** |
| Value | % | This group includes non-COVID-19 medical personnel (doctors, nurses, and other therapists) in permanent contact with non-COVID-19 patients in medical institutions. Risk factor evaluation rating scale:<br>0 = no risk, 1 = low, 2 = medium, 3 = high, 4 = very high |
| 0 | 0 |  |
| 4 | 100 |  |
| **12212** |  | **Non-medical personnel in medical institutions** |
| Value | % | This group includes non-medical personnel (administrators, info-desk, security, parking, lab personnel) in intermittent (usually short) contact with patients and/or medical personnel in medical institutions. Risk factor evaluation rating scale:<br>0 = no risk, 1 = low, 2 = medium, 3 = high, 4 = very high |
| 0 | 0 |  |
| 4 | 100 |  |
| **1222** |  | **Non-medical service personnel risk factors** |
| Value | % | This group includes all non-medical professions that require permanent contact with customers (cashiers, restaurant workers, taxi drivers, hair stylists, flight attendants, airport personnel, F2F teachers, bank tellers clerks, etc.). Risk is evaluated using the following 7-level scale:<br>0=none, 1 = very low, 2 = low, 3 = medium, 4 = high, 5 = very high, 6 = extreme |
| 0 | 0 |  |
| 6 | 100 |  |
| **1223** |  | **Essential emergency personnel risk factors** |
| Value | % | This group includes firefighter, military, and security personnel in low exposure activities. Risk factor evaluation rating scale:<br>0 = no risk, 1 = low, 2 = medium, 3 = high, 4 = very high |
| 0 | 0 |  |
| 4 | 100 |  |
| **1224** |  | **Other professions risk factors** |
| Value | % | This group includes all professions not included in previous criteria. No risk or low risk rating is assigned to people who do all work from home or people in low exposure activities (e.g. farmers, rangers, online instructors, etc.) Risk factor evaluation rating scale:<br>0 = no risk, 1 = low, 2 = medium, 3 = high, 4 = very high |
| 0 | 0 |  |
| 4 | 100 |  |
| **12311** |  | **Retirement and nursing homes** |
| Value | % | Retirement homes and assisted living institutions create risks based on proximity and age of patients. This criterion also applies to retirement home service personnel. Evaluation is based on the following risk rating scale:<br>0 = no risk, 1 = low, 2 = medium, 3 = high, 4 = very high |
| 0 | 0 |  |
| 4 | 100 |  |
| **12312** |  | **Non-retirement home options** |
| Value | % | Evaluated using the following (or similar) group living options:<br>(1) Life in family, group homes, or homeless shelters with n members:<br>Score = $100 \cdot \min[1, (n-1)/5]$<br>(2) Student dorm: 0 = no risk, 25 = low, 50 = medium, 75 = high, 100 = very high<br>(3) Prison: 0 = no risk, 25 = low, 50 = medium, 75 = high, 100 = very high |
| 0 | 0 |  |
| 100 | 100 |  |
| **12321** |  | **Degree of urbanity** |
| Value | % | High degree of urbanity increases risks of COVID-19 transmission. Evaluated using the following rating scale:<br>0 = very low (isolated rural areas), 1 = low, 2 = medium, 3 = high, 4 = very high |
| 0 | 0 |  |
| 4 | 100 |  |
| **12322** |  | **Density of population** |
| Value | % | High density of population increases contacts between people and facilitates the spread of COVID-19. Evaluated using the following rating scale:<br>0 = very low, 1 = low, 2 = medium, 3 = high, 4 = very high |
| 0 | 0 |  |
| 4 | 100 |  |

| 131 |  | Disadvantaged groups |
| --- | --- | --- |
| Value | % | This criterion includes all ethnic/social/racial minority groups disadvantaged in relation to COVID-19 vaccination and health care (based on religion, place of residence, race, ethnicity, education, occupation, etc.). According to CDC that includes persons from correctional facilities, homeless shelters, people with intellectual or development disabilities, substance use disorder, and sexual and gender minorities. Risk evaluation rating scale:<br>0 = no risk, 1 = low, 2 = medium, 3 = high, 4 = very high |
| 0 | 0 |  |
| 4 | 100 |  |
| 132 |  | Economic status |
| Value | % | This group includes people living in poverty with poor access to health care. People in this group usually experience also the problem in necessary time and travel cost to go to places offering a free vaccine.<br>Risk evaluation rating scale: 0=no risk, 1=low, 2=medium, 3=high, 4=very high |
| 0 | 0 |  |
| 4 | 100 |  |
| 133 |  | Social, educational, cultural, and family factors |
| Value | % | This group includes people with insufficient education, low socioeconomic status, family problems (no parents, single parents, family conflicts), cultural and language isolation, and similar factors that reduce health care equity. COVID-19 risks for people in this group are evaluated using the following rating scale: 0 = no risk, 1 = low, 2 = medium, 3 = high, 4 = very high |
| 0 | 0 |  |
| 4 | 100 |  |

In most of the presented criteria the arguments don’t have measurable numeric values. For example, the current health conditions (#12121) cannot be directly measured like age, but they can be evaluated using a rating scale (excellent, good, average, poor, very poor), and primary care providers can easily provide such evaluation for their patients. In most cases it is sufficient to have a 5-level scale. If it is desirable and possible to provide higher precision the rating scales can have more levels; e.g., the non-medical personnel risk factors (#1222) use a 7-level scale.

The attribute criteria reflect the recommendations provided in [1-8]. They can be modified, expanded or reduced according to the situation in a given community, but in all cases they should be used to obtain all attribute priority degrees *p*_1_, ⋯, *p*_*m*_ as individual contributors to the resulting vaccination priority degree. The resulting vaccination priority degree is obtained using a compound aggregation function of *m* contributing inputs. The proposed LSP criterion and all numeric results in this paper are generated using the LSP.NT decision support tool [14].

## 5. Logic aggregation

The computation of the overall priority *P* = *L*(*p*_1_, ⋯, *p*_*m*_) is based of functions that can model various logic requirements that the inputs must satisfy;

- Adjustable degree of simultaneity
- Adjustable degree of substitutability
- Mandatory requirements (all inputs must be at least partially satisfied)
- Sufficient requirements (a single fully satisfied input is sufficient to (almost) completely satisfy a group of requirements)
- Adjustable degrees of importance of inputs
- Optional inputs (all inputs are desirable but neither mandatory, nor sufficient)
- Mandatory/optional inputs (combination of mandatory inputs and optional inputs)
- Sufficient/optional inputs (combination of sufficient inputs and optional inputs)

All priority attributes individually and independently contribute to the vaccination priority, and no one of them depends on other attributes in its group. In addition, many of inputs are mutually exclusive: if one is satisfied then others cannot be satisfied (e.g., if an individual works in a medical profession, s/he is not working in non-medical professions, and vice versa). This is a rather rare situation in evaluation where the whole aggregation structure is disjunctive. In this case aggregation can be based on simple weighted power means 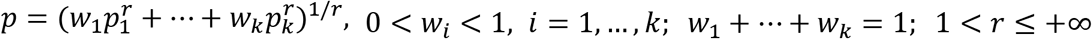. The weights *w*_*i*_ denote the degree of importance of input *p*_*i*_ and the exponent *r* is used to adjust the degree of disjunction (orness [13]) in equidistant steps from the lowest to the highest. If the orness step is 1/16, then the adjustable partial disjunctive aggregators are located between the arithmetic mean A and the pure disjunction D, and denoted as follows: A, D--, D-, D-+, DA, D+-, D+, D++, D. Here D-, DA and D+ denote respectively a weak, average, and strong partial disjunction; they can be made stronger or weaker, and that is the role of aggregators D--, D-+, D+-, and D++. The strongest disjunctive aggregator D, obtained using *r* = +∞, denotes the pure disjunction, i.e. the function *max*(*p*_1_, ⋯, *p*_*k*_). These aggregators are used in the aggregation structure shown in Fig. 4 (more details about LSP aggregation operators can be found in [13]).

**Fig. 4.**
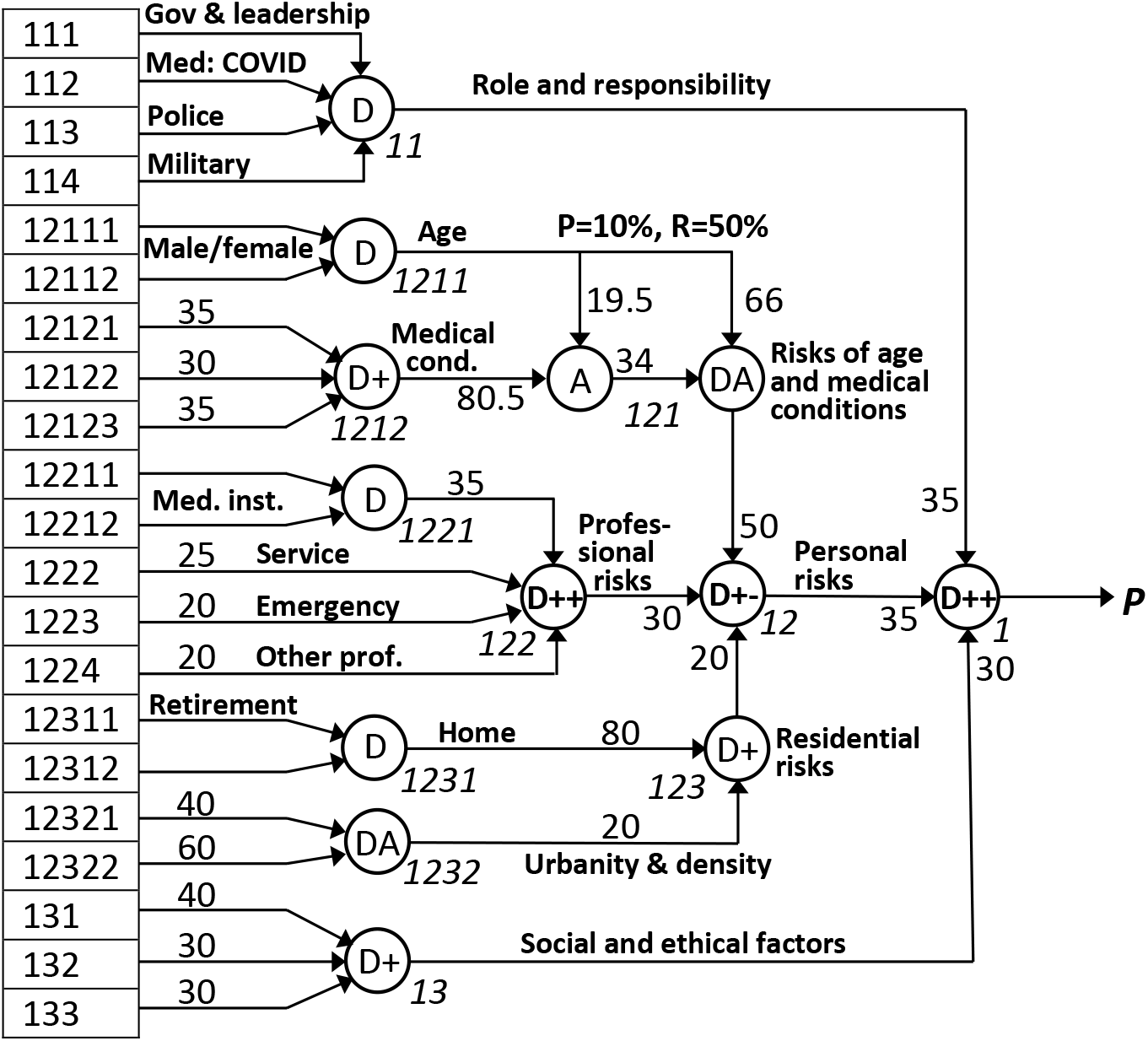
Logic aggregation structure for computing COVID-19 vaccination priority degree

The aggregation structure shown in Fig. 4 includes four aggregators that aggregate mutually exclusive inputs: unique and different social/professional roles (#11), male/female age (#1211), retirement/non-retirement home (#1221), and medical/non-medical personnel (#1231). In all these cases there is only one nonzero input justifying the use of the pure disjunction. In all other cases, all inputs can contribute to output values and it is appropriate to use weighted partial disjunction aggregators. For example, the personal medical conditions (#1212) can be simultaneously strongly affected by current conditions, medical history, and race and ethnicity; that justifies the use of strong partial disjunction.

The aggregator #121 (a combination of aggregators A and DA) is called the disjunctive partial absorption. According to all governmental recommendations, age is a primary and sufficient reason for high priority. Consequently, the age input alone is considered sufficient to cause the high priority. On the other hand, the high priority can also be assigned to younger people, provided that their personal medical conditions are not good and provably amplify risk factors. The risk factors *F*(*a, m*) are the function of age *a* and medical conditions *m*. For given age we assume the following properties: *F*(*a*, 0) = *a* − *p*_0_(*a*), *F*(*a*, 1) = *a* + *r*_1_(*a*), where *p*_0_(*a*) is called penalty and *r*_1_(*a*) is called reward. In Fig. 4, the mean value of *p*_0_(*a*) is denoted *P* and called the mean penalty and the mean value of *r*_1_(*a*) is denoted *R* and called the mean reward. The properties of the partial absorption aggregator are adjusted by selecting desired values of *P* and *R*. In our example in Fig. 4, we selected a small penalty *P*=10 % to show the dominant impact of age, and the significant reward of 50% to show that poor medical conditions can significantly affect the vaccination priority even for relatively young people.

## 6. Examples and discussion

The proposed LSP criterion can be validated using a set of individuals that have properties characteristic for four phases initially presented in [5] and subsequently refined in [6-8]. Fig. 5 shows the sample population that we use for the LSP criterion validation using LSP.NT [14]. Detailed results of evaluation are presented in Fig. 6.

**Fig. 5.**
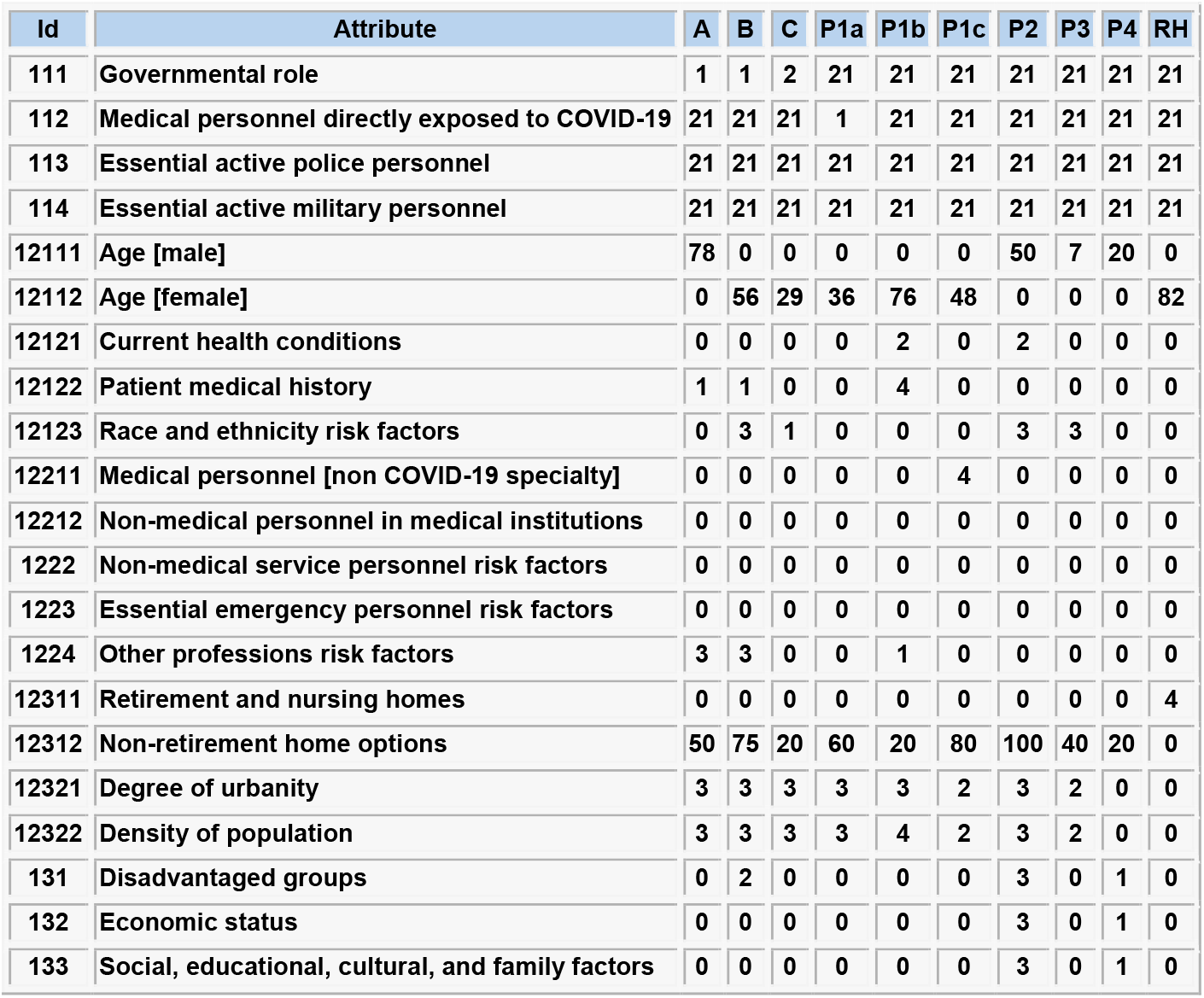
Attribute values for the sample population of nine people

**Fig. 6.**
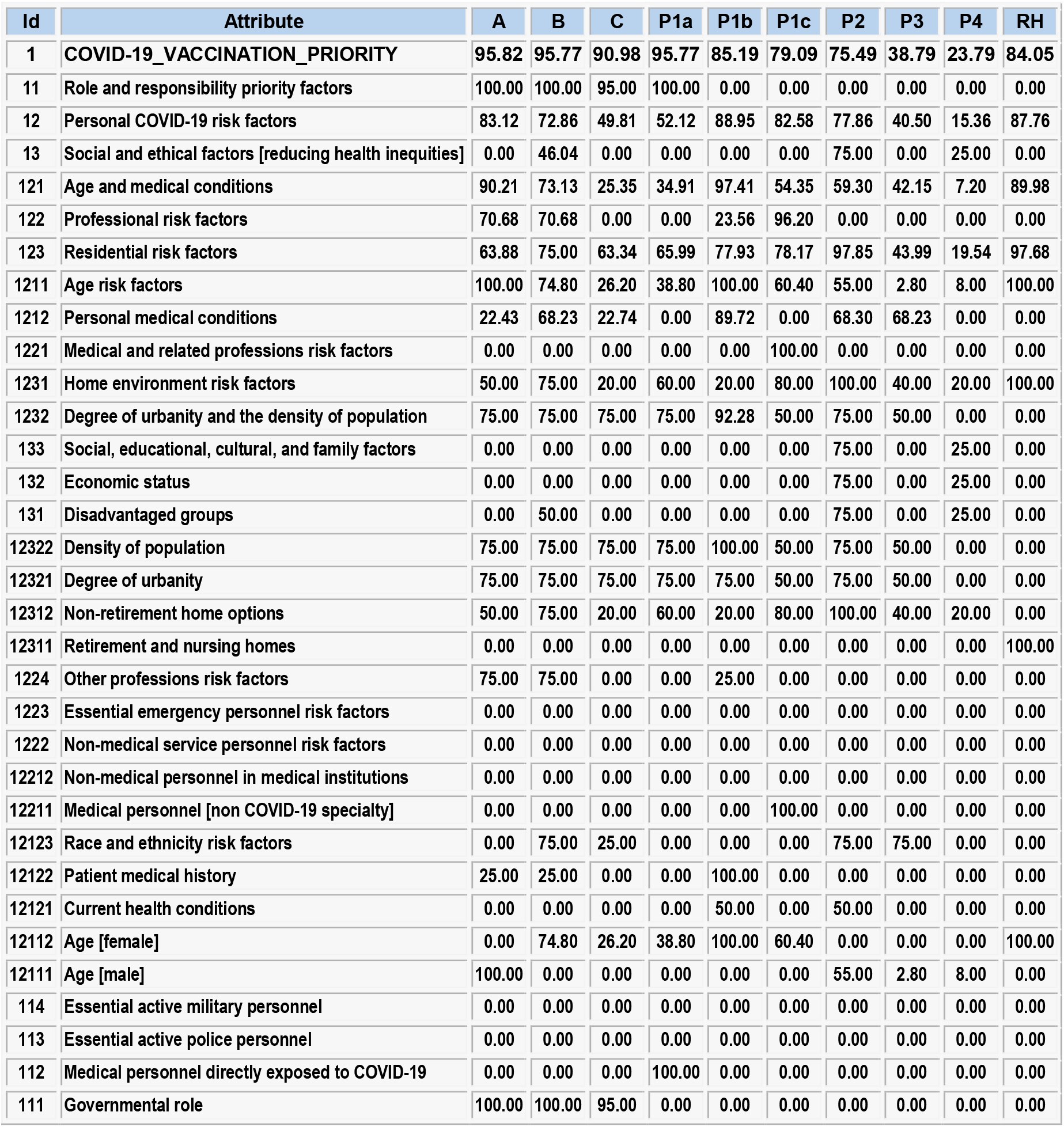
Results of the vaccination priority evaluation for the sample population

Ten individuals included in this evaluation are defined as follows:

**A** = President of the United States

**B** = Vice president of the United States

**C** = Congresswoman, 29 years old, healthy, family of 2, living in highly urban area

**P1a** = Medical doctor or nurse in permanent contact with COVID-19 patients; female, 36 years old, in good health, living with husband and two children in a dense highly urban area

**P1b** = Woman, 76 years old, cancer survivor, in average current health conditions, living with husband in a dense highly urban area

**P1c** = Obstetrician, female, 48 years old, excellent health, living with husband and three children in a moderately dense urban area

**P2** = Man, 50 years old, average health conditions, high race risk factor, living in a homeless shelter, high risks in all social and ethical categories.

**P3** = Child, 7 years old, high ethnicity risk factors, living with parents in an average urban area.

**P4** = A healthy farm worker, 20 years old, family of 2, low risk in all social and ethical categories.

**RH** = Woman, 82 years old, average health, living in a high risk retirement home (long-term care).

The obtained COVID-19 vaccination priority results, shown in the first row of the table in Fig. 6, approximately correspond to expected values recommended in [1-9]. It is very easy to modify and adjust parameters of the proposed criterion to make it more precise representation of specific medical and/or social requirements.

In the criterion adjustment process it is useful to perform the sensitivity analysis exemplified in Fig. 7. If the age of the hypothetical healthy farm worker P4 increases from his current age 20, then the vaccination priority would also increase in the range from 23.79% to 77.68%. If these values need to be modified, the LSP criterion can be adjusted to provide desired different results.

**Fig. 7.**
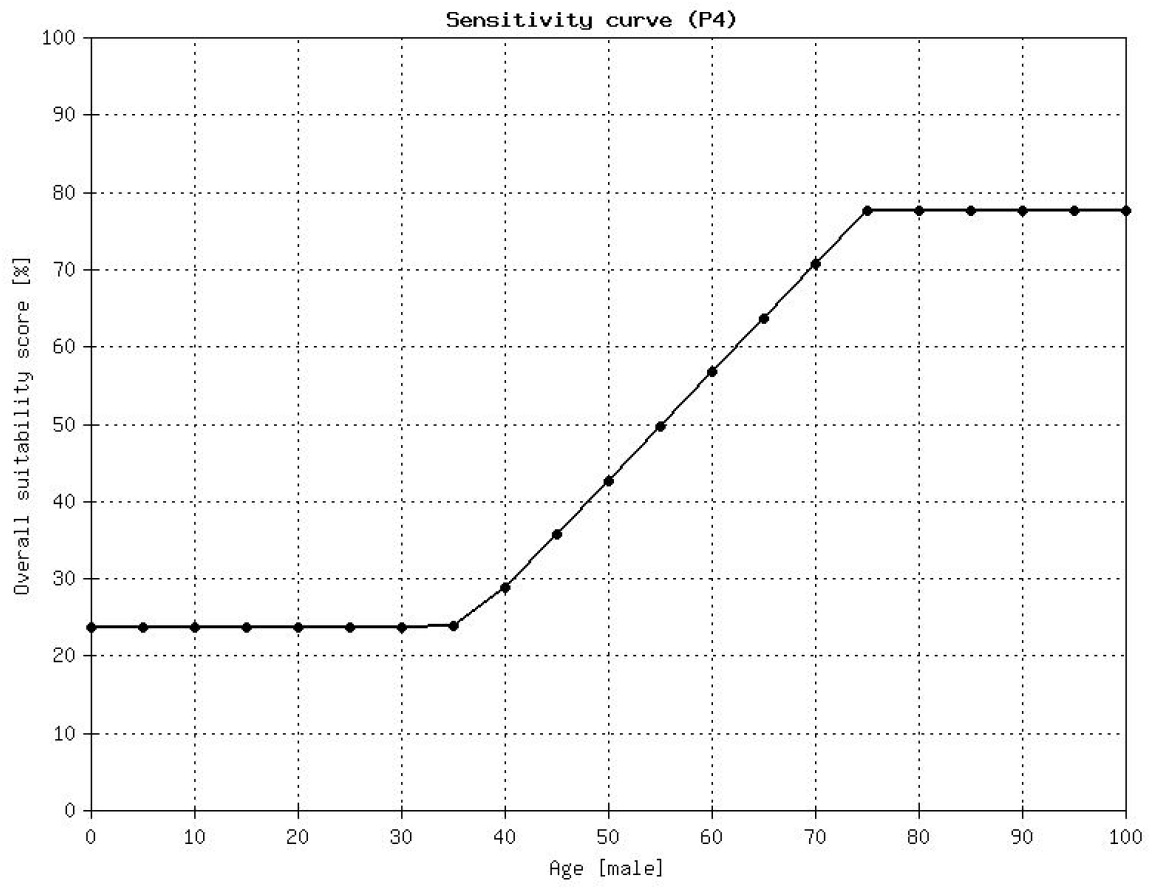
Sensitivity analysis for the age of the farm worker P4

Practical use of LSP criteria depends on the availability of input attributes. Some of them can be obtained automatically from available databases (e.g. age, and the degree of urbanity based on patient’s address), some of them must be provided by the patient (e.g. current employment, number of people in the household, race, ethnicity, etc.), and some inputs can only be provided by a medical professional. The LSP method is tolerant to missing data [13, 14] and the vaccination priority can be computed even in cases of incomplete input attribute data.

## 7. Conclusions

COVID-19 vaccination priority is a decision problem that can and should be solved using quantitative decision support methods. Our solution is based on the LSP method and includes 21 input attributes. The fundamental goal of the proposed criterion is to be complete: it includes all types of inputs: social, medical, and ethical. While the presented default version of the LSP criterion is based on governmental documents and recommendations, it is easy to adjust the criterion function (attribute criteria, aggregation operators and their weights) in a way that better reflects specific requirements of a selected region or community. In addition, the presented evaluation methodology can be used in many other medical priority evaluation problems, and in particular, in the organ transplant priority decision making.

In the case of the COVID-19 priority vaccination problem, the benefits obtained using the quantitative LSP evaluation are obvious: better planning and organization of vaccine distribution, the reduction of mortality from COVID-19, the reduction of expenses and the wasted time of all involved, and a political benefit of more confident and satisfied vaccinated population.

## Data Availability

The manuscript includes a full disclosure of all data we used in our examples.

http://www.seas.com/LSPNT/login.php

